# Leveraging Large Language Models to Identify In-Hospital Cardiac Arrest

**DOI:** 10.1101/2025.10.16.25338028

**Authors:** Jonathan Vo, Davy Weissenbacher, Kyndaron Reinier, Harpriya Chugh, Audrey Uy-Evanado, Karen O’Connor, Graciela Gonzalez-Hernandez, Sumeet S. Chugh

## Abstract

Manual chart abstraction is the gold standard for identifying in-hospital cardiac arrest (IHCA) but is resource-intensive. Diagnosis codes are a widely used alternative given their accessibility and automated nature, but this method has poor sensitivity and positive predictive value. We present a novel large language model (LLM) approach to identify IHCA and location, highlighting the potential of LLMs for rapid, accurate, and automated IHCA identification from clinical notes.

---

In-hospital cardiac arrest (IHCA) affects approximately 300,000 patients in the United States annually, representing 1.6 to 2.8 patients per 1000 hospital admissions ^21^. IHCA is also associated with a high mortality rate, estimated at least 75% ^4^. The standard definition of IHCA is cessation of cardiac activity requiring chest compressions and/or defibrillation (also known as cardiopulmonary resuscitation or CPR) among patients admitted to the hospital ^12^.

Owing to the high morbidity and mortality of IHCA as well as its rising incidence ^19^, efforts to identify predictive factors and improve treatment have contributed to improvements in survival. However, the survival rate from IHCA has plateaued in recent years ^21^ signaling a need for increased research efforts and high quality cardiac arrest registries to improve resuscitation quality and outcomes. This goal is highlighted as “step two” of the American Heart Association’s Ten Steps Toward Improving IHCA Quality of Care and Outcomes, which emphasizes collection of high quality data to improve resuscitation care ^10^.

Much of current IHCA research utilizes large prospective registries which require trained personnel to abstract medical records and cardiac arrest flow sheets. The largest of these registries in the US is the Get With The Guidelines – Resuscitation (GWTG) registry developed by the American Heart Association ^14^. However, only 5-6% of US hospitals participate in this registry in any given year leaving the majority of this research resource untapped ^18^.

For hospitals that do not participate in such research registries, cases are identified in a prospective manner using the gold standard of manual chart abstraction, but these are labor-intensive methods requiring trained personnel. Alternatively, research studies identify IHCA in a retrospective manner often using International Classification of Disease (ICD) diagnosis codes for quick ascertainment of research populations from the electronic health record (EHR). However, ICD codes alone have been found to have low positive predictive values (PPV) and low sensitivity for identifying IHCA cases ^2;6^. This may be due to a number of reasons including misclassification or inaccurate coding of diagnoses, ICD codes referring to an earlier encounter, and inappropriate omission of ICD codes due to human error. Regional variability and inconsistency in ICD coding across different longitudinal time points further limits usability of ICD codes for research studies ^11^.

Some previous studies have used natural language processing (NLP) and large language models (LLM’s) to automatically identify individuals with other conditions such as heart failure, recurrent atrial fibrillation, and heart disease risk factors from raw clinical text; however, no such work has been done for IHCA case detection ^1;5;8;22^, especially since the nuance of location and timing of the event is challenging.

In this study, we utilized two distinct data sets. The first set, the *Evaluation Corpus*, comprised of a mix of 500 Cedars-Sinai Medical Center EHR encounters; 47% had an ICD code indicating IHCA and 53% were randomly selected inpatient encounters. The ICD codes used included I49.01, 49.02, 46.2, 46.8, 46.9, CPT code 92950, and procedure codes 5A12012 and 02QA0ZZ. An encounter was defined as all clinical notes for one hospital visit. Approximately half of the Evaluation corpus included ICD codes for IHCA to ensure enough positive IHCA cases for our system. Two physicians (JV and AUE) annotated the Evaluation Corpus for adjudication of IHCA and location with this manual annotation utilized as the gold standard. The determination of IHCA was made according to the widely accepted Utstein definition: cardiac arrest of a patient admitted to Cedars-Sinai Medical Center requiring chest compressions and/or defibrillation ^12^. For the purposes of our study, location was categorized as 1) In-hospital (IH) including critical care units, operating rooms, and cardiac catheterization laboratories, 2) emergency department (ED), and 3) out-of-hospital cardiac arrest for which treatment was initiated out of the hospital and continued on arrival to the emergency department (OH-ED). The third category OH-ED (Out-of-hospital cardiac arrest with continued treatment in the ED), is traditionally not part of standard IHCA definitions but we included it to facilitate identification of this sub-group and enable their potential exclusion in future research. The Evaluation Corpus was randomly split into training (300 encounters, 60%), development (50 encounters, 10%), and testing (150 encounters, 30%) sets. Two physicians independently double-annotated 50 encounters for presence and location of IHCA and achieved a Cohen’s kappa score of 0.951 indicating almost perfect agreement for the annotation of IHCA presence, and a Cohen’s kappa score of 0.838 indicating strong agreement for IHCA location annotation ^9^.

We used a second data set, the *Observational Study of Sudden Cardiac Arrest (OSCAR) corpus*, that contained 45,525 encounters from an ongoing observational study of patients who received care at Cedars Sinai Health System. This cohort has been described previously ^15^. These encounters included all inpatient, ED, or observation visits as well as any encounter with a code blue note; no ambulatory encounters were included. All patient encounters during the year 2023 were included without any pre-selection or exclusion; therefore, the prevalence of IHCA within the OSCAR corpus is closer to the general population.

We deployed a LLM as a multiclass classifier. Specifically, we prompted the LLM to identify IHCA cases in EHR clinical notes and determine their broad location (i.e. IH, ED, OH-ED), or assign ‘None’ to notes without an IHCA mention. At the encounter level, we labeled an encounter as positive if at least one of the encounter notes mentioned IHCA. We then defined the location of the encounter-level IHCA as the location of the first IHCA event in the encounter note. To develop our system, we began with a basic prompt that simply instructed the LLM to identify notes that listed a cardiac arrest and its location. We then progressively refined the prompt –first adding conditional guidelines, then providing few-shot examples with labels, followed by instructing the LLM to include reasoning in its outputs and illustrating this reasoning within the same few-shot examples, and finally incorporating a self-correction step. We improved the prompt using the training and development sets and evaluated the final performance of the LLM on the test set.

Using our Evaluation corpus, we evaluated the binary performance of the LLM determining whether a clinical encounter mentioned an IHCA irrespective of its location - using standard metrics: precision, recall, and F1 score. We assessed multiclass classification performance with a confusion matrix ^17^. Our metrics evaluated encounter-level predictions as these are more relevant in clinical contexts. Because precision varies with disease prevalence (as opposed to recall which does not), we further evaluated the LLM’s performance on the OSCAR corpus using precision alone. For this purpose, two adjudicators (AUE and JV) manually adjudicated all of the positive IHCA predictions made by the LLM and achieved a Cohen’s kappa score of 0.89 indicating high agreement between the adjudicators. This manual adjudication was used as the gold standard. We compared our system against three baselines: (1) the LLM with the final prompt but without self-correction, (2) the LLM with only the basic prompt, and (3) a conventional ICD-based algorithm.

Our system achieved high performance on the test set of the Evaluation corpus with a precision of 0.88, recall of 0.97, and F1 score of 0.92 (Table 1). In comparison, the ICD-code baseline performed substantially worse, reaching a precision of 0.68, recall of 0.91, and F1 score of 0.78. While the LLM missed a small number of encounters that were manually determined to be an IHCA by our gold standard two-physician adjudication, it more often generated false positives by predicting IHCA mentions in encounters were not adjudicated as IHCA. As seen in Figure 1, the LLM also accurately identified location of IHCA in most cases with majority of errors occurring in predicting IH with cases of no IHCA, predicting ED arrest in cases of no IHCA, and predicting IH arrest in cases of ED arrest.

**Table 1.** Encounter-level IHCA detection performance on the Evaluation and OSCAR Corpus.

|  | Evaluation corpus |  |  | OSCAR corpus |
| --- | --- | --- | --- | --- |
|  | Prec. | Rec. | F1 | Prec. |
| Final Prompt + Self-Correction | 0.88 | 0.97 | 0.92 | 0.79 |
| Final Prompt | 0.86 | 1.0 | 0.93 | 0.47 |
| Basic Prompt | 0.77 | 1.0 | 0.87 | — |
| ICD Codes | 0.68 | 0.91 | 0.78 | — |

Precision, Recall, and F1 scores of our system on the test set of the Evaluation corpus (150 patient encounters) and the OSCAR corpus, looking at encounter-level predictions. Basic prompt includes giving the role and instructions. Final prompt includes the basic prompt along with conditional guidelines, chain-of-thought, and few shot learning.

**Fig. 1.**
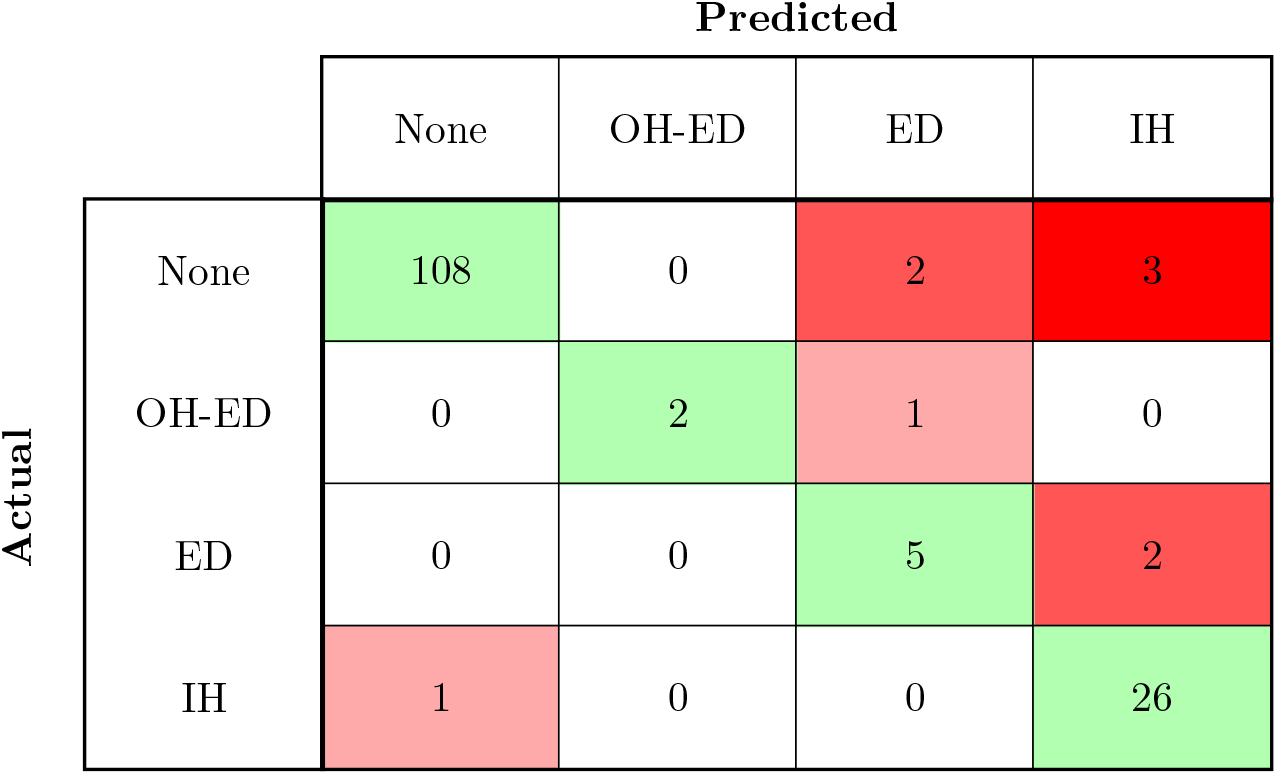
Confusion matrix for the prediction of our system on the test set of the Evaluation corpus. Values along the diagonal are correct encounter-level IHCA predictions. Values in red are incorrect predictions. None refers to no IHCA. OH-ED refers to SCA that begins outside of the hospital and with treatment continuing in the ED. ED refers to arrest that begins in the ED. IH refers to arrest that begins while admitted to the hospital.

Performance of the LLM on the OSCAR corpus remained high with a precision of 0.79 (Table 1). Its lower precision on this data compared to the test set of the Evaluation corpus likely reflects to the lower prevalence of IHCA in this corpus. The LLM accurately identified encounter-level IHCA location in most cases as well. As seen in Figure 2, errors in the OSCAR corpus were similar to the Evaluation corpus. Again, most errors occurred in predicting IHCA in cases of no IHCA, predicting ED arrest in cases of no IHCA, and predicting IH arrest in cases of ED arrest.

**Fig. 2.**
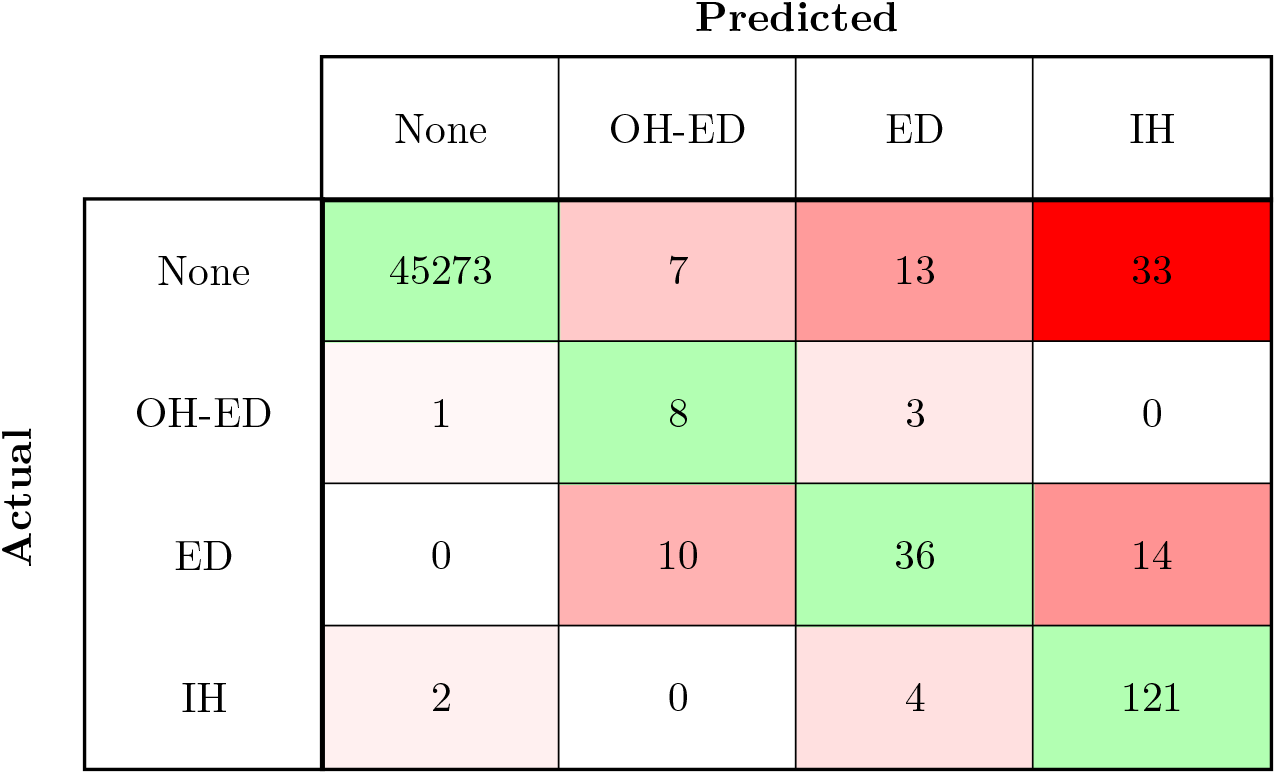
Confusion matrix for OSCAR corpus. Values along the diagonal are correct encounter-level IHCA predictions. Values in red are incorrect predictions. None refers to no IHCA. OH-ED refers to SCA that begins outside of the hospital and with treatment continuing in the ED. ED refers to arrest that begins in the ED. IH refers to arrest that begins while admitted to the hospital.

We also carefully examined the types of errors made by the LLM. Errors included mention of an arrest but no treatment, human error of annotation, incorrectly categorizing unstable rhythms with rapid pulses as a cardiac arrest, and incorrect classification of location.

In conclusion, our study demonstrates the feasibility of using a publicly available LLM to identify IHCA events and their locations in unfiltered clinical encounters, highlighting their potential to support real-world clinical research. To the best of our knowledge, this is the first study to apply an LLM for identification of IHCA events at this level of detail. Even with a basic prompt, our system outperformed a conventional ICD-code algorithm, and further prompt engineering yielded additional improvements. These findings suggest that LLMs could provide higher-fidelity, lower-cost alternatives to ICD codes and manual adjudication for automated IHCA identification.

At the same time, our work underscores several key opportunities for systematic evaluation and improvement. The model was developed and evaluated on a single health system’s EHR, and external validation will be essential to demonstrate generalizability. We also focused on basic prompt engineering to improve the system; future work should explore more advanced strategies, such as searching similar notes for few-shot prompting, fine-tuning the LLM with task-specific supervision and/or reinforcement learning with human feedback ^13^. Moreover, most publicly available LLMs are trained primarily on general internet data with limited exposure to clinical text, which is protected for privacy reasons, limiting their knowledge and applicability in clinical settings. ^16^ We partially mitigated this by providing clinical guidelines and precise definitions in lay terms, but even greater gains may come from models pre-trained on large amount of medical documents and instructed to follow domain-specific reasoning ^7^. Finally, despite our prompt engineering efforts, the model still produced a relatively high number of false positives, which would preclude its direct use in current research studies. A two-pass system — using the LLM for high-recall initial filtering followed by physician adjudication — holds promise for efficiently creating high-quality IHCA registries while reducing manual effort.

## Methods

### Evaluation Corpus Collection and Annotations

The study was approved by the Cedars-Sinai Medical Center Institutional Review Board. We evaluated our system on 500 clinical encounters, which we collected from ICD-coded cases (235 encounters) and randomly selected inpatient encounters (265 encounters). Because cardiac arrest is rare, we expected most random cases to be negatives; therefore, we included ICD-coded cases to ensure sufficient true positives for training and testing. The ICD codes used are shown in Table 2.

**Table 2.** ICD/CPT Codes and Corresponding Descriptions for Cardiac Arrest.

| ICD/CPT Code | Description |
| --- | --- |
| I40.01 | Ventricular fibrillation |
| I40.02 | Ventricular flutter |
| I46.2 | Cardiac arrest due to underlying cardiac condition |
| I46.8 | Cardiac arrest due to other underlying condition |
| I46.9 | Cardiac arrest, cause unspecified |
| 92950 | CPT code for CPR (procedure code) |
| 5A12012 | Procedure for CPR: "Performance of Cardiac Output, Single, Manual" |
| 02QA0ZZ | Procedure for CPR: "Repair Heart, Open Approach" |

A clinical encounter encompassed one hospital stay for a patient, so one patient may have multiple clinical encounters. Clinical encounters included all inpatient notes for during the encounter dates such as emergency department (ED) notes, discharge summary, as well as any Code Blue notes which are notes written after attempted resuscitation of an IHCA. One clinical encounter may include one or up to all three note types. There may be multiple notes of the same note type within one encounter.

We defined IHCA according to the widely accepted Utstein definition: cardiac arrest of an admitted patientrequiring chest compressions and/or defibrillation. Two physicians annotated the 3,822 notes corresponding to the 500 clinical encounters. They labeled the notes as *IHCA* if the clinical text provided clear evidence that a patient met this definition, otherwise as *not IHCA*. Before beginning annotation, we developed a set of guidelines through an iterative process. The two annotators independently reviewed a pilot sample of 50 notes and adjudicated any disagreements, revising the guidelines as necessary before repeating the process. Iterations continued until no further modifications were required. This approach ensured consistency across annotations and enhanced the reproducibility of our study on other corpora. The full set of guidelines used by annotators for determination of IHCA are shown in Table 3.

**Table 3.** Guidelines used by annotators to assess for presence of IHCA. Criteria for IHCA was based on the standard Utstein definition of pulselessness that is treated with chest compressions or defibrillation in the hospital.

| **Criteria for IHCA:** An encounter was positive for IHCA if both of the following conditions are met:<br>1. The patient became pulseless<br>2. Due to this pulselessness, the patient required treatment with chest compressions and/or external defibrillation |  |
| --- | --- |
| Rule | Description |
| **Rule 1 (Positive)** | If the patient became pulseless requiring treatment with chest compressions and/or external defibrillation while admitted to the hospital, then the patient experienced in-hospital cardiac arrest (IH). |
| **Rule 2 (Positive)** | If the patient became pulseless requiring treatment with chest compressions and/or external defibrillation in the emergency department, then the patient experienced in-hospital cardiac arrest located in the emergency department (ED). |
| **Rule 3 (Positive)** | If the patient became pulseless requiring chest compressions and/or external defibrillation outside of the hospital, but these measures were continued on arrival to the hospital, then the patient experienced in-hospital cardiac arrest that started out of the hospital (OH-ED). |

Table 3 – continued from previous page
| Rule | Description |
| --- | --- |
| **Rule 4 (Positive)** | If a patient had pre-existing do-not-resuscitate orders and became pulseless but inadvertently had treatment with chest compressions and/or external defibrillation in the hospital or emergency department, then the patient experienced in-hospital cardiac arrest. |
| **Rule 5 (Negative)** | If a patient received chest compressions and/or external defibrillation but did not lose pulses, then the patient did not experience in-hospital cardiac arrest. |
| **Rule 6 (Negative)** | If patient was on comfort care or do-not-resuscitate (DNR/DNAR) and had a cardiopulmonary arrest, then the patient did not experience in-hospital cardiac arrest. |

For each note that we labeled as positive for IHCA, we also labeled the location of the arrest. Possible locations of IHCA for this study were in-hospital (IH), emergency department (ED), or out-of-hospital with treatment continuing in the emergency department (OH-ED). For notes documenting multiple arrests, we only labeled the mentions of the first arrest. As we were mainly interested in in-hospital cardiac arrest, we did not include cases of only out-of-hospital arrest with successful resuscitation prior to arrival to the hospital. OH-ED cardiac arrests are traditionally as OHCA; however, these were included in this study to be treated similarly to ED arrests and for potential exclusion of these cases in future research studies using our system.

Following annotation of each individual note within an encounter, two physicians reviewed the entire clinical encounter comprising of all notes together to annotate the encounter’s IHCA adjudication and location as a whole. If there were multiple IHCA events, location of the encounter was taken from the first event. As an example, if there was evidence of IHCA within the discharge summary but not the ED note, the encounter would be annotated as positive for IHCA, and the discharge summary would also be annotated as positive for IHCA, but the ED note would be labeled as negative for IHCA.

We randomly split the 500 encounters into a training set (300 encounters, 60%), development set (50 encounters, 10%), and test set (150 encounters, 30%). The training set included 99 encounters with at least one IHCA, comprising 172 positive notes. The development contained 11 positive encounters with 22 positive notes, and the test set 37 positive encounters with 70 positive notes. Positive encounters and notes were determined by manual annotation as described above.

Because the Evaluation corpus had a high prevalence of IHCA due to it consisting of a mix of potential IHCA cases by ICD codes as well as random clinical encounters, we validated the performance of the LLM on a corpus of encounters obtained between the dates 1/1/2023 and 12/31/2023 from the ongoing Observational Study of Sudden Cardiac Arrest (OSCAR) cohort. This cohort has been described previously ^15^. These encounters included all inpatient, ED, or observation visits or anyone with a code blue note(total n=45,525) between those dates and allowed us to determine the LLM’s precision on data with a prevalence of IHCA closer to the general population. Positive IHCA predictions of this corpus were adjudicated by two physicians (JV and AUE) to identify true positive and false positive predictions by the gold standard of manual review. Adjudication was performed by two physicians who reviewed all inpatient notes within a clinical encounter and identified if the encounter met the standard Utstein definition of IHCA(pulselessness requiring treatment with chest compressions and/or defibrillation).

### LLM choice and Prompt Development

In recent years, several large language models have been released to the research community. We selected Meta’s LLaMA 3.1-405B pre-trained and instruction-tuned generative model because it is open-source and its scale is sufficient to deliver state-of-the-art performance while remaining computationally feasible for inference ^3^.

We compared the performance of four systems. In the first baseline, we instructed the LLM by specifying its role, providing a concise description of the task, i.e. classifying notes mentioning IHCAs and their locations, and appending the note to be classified.

In the second baseline, we refined the prompt by supplying the LLM with additional task-specific information. We first augmented the initial prompt with a summary of the clinical guidelines that physicians followed during the annotation of the notes. Because the Llama 3.1-405B model was pretrained on publicly available online data, it had minimal exposure to medical documents authored by professional clinicians, which are generally inaccessible due to privacy protections. This limitation reduced the model’s familiarity with medical knowledge and terminology. To address this gap, we added to our prompt concise definitions of all relevant concepts required for the task, and reformulated the annotation guidelines into a set of clear, implication-based rules.

Following standard prompt engineering practices, we also employed few-shot learning combined with chain-of-thought prompting to further enhance the prompt. We implemented few-shot learning by adding a small set of manually selected examples of notes with their correct labels (IHCA +/-location) from the training set to guide the LLM’s predictions. During several iterations of error analysis on the training set, we identified common errors made by the LLM and curated representative examples to include in the prompt. Most errors were false positives, but we also included a few true positive and true negative cases. Using chain-of-thought prompting, we asked the LLM to briefly explain its predictions and support its claims with direct inline quotations from the notes. Encouraging the LLM to generate step-by-step reasoning helps reduce hallucinations and improves output reliability. ^20^

We built our final system on the second baseline, adding a self-correction step. Using the same initial prompt as the baseline, we first obtained the classification and reasoning, then had the LLM re-evaluate its output. Since most errors were false positives or incorrect location assignments, we restricted this review to notes initially classified as positive IHCA encounters. If the model predicted a note as negative, we moved directly to the next note. For positive predictions, we passed the classification, location, reasoning, and note text to a secondary prompt instructing the model to confirm or revise its decision. We created three such prompts, one for each location category: IH, ED, and OH-ED.

#### Testing and Validation

We evaluated the performance of each baseline and our final system on the test set of our Evaluation corpus. We assessed the LLM sequentially: first with the initial prompt, then with the final prompt (the initial prompt augmented with clinical guidelines, few-shot examples, and chain-of-thought prompting) and finally with the final prompt plus self-correction. We specifically looked at encounter-level predictions rather than individual note-level predictions as the encounter predictions are more relevant for clinical and research purposes since it matters more whether a patient has experienced IHCA at all rather than if an individual note mentions IHCA. Additionally, ICD codes identify IHCA at the encounter level, so this allowed us to compare our LLM approach to ICD codes.

We compared the performance of the LLM to ICD codes that are commonly used to identify IHCA which include ICD codes I49.01, 49.02, 46.2, 46.8, 46.9, CPT code 92950, and procedure codes 5A12012 and 02QA0ZZ. Testing of the ICD codes was done in a two-step manner. First, to test for recall of the ICD codes, we used the above ICD codes to identify all cases of IHCA from a gold standard list of patients with known IHCA. This gold standard list was all encounters in the annotated corpus that were annotated as positive. Cases missed by the ICD codes were flagged as false negatives. Next, to test for precision of the ICD codes, we identified potential cases of IHCA from a cohort of patients within the dates 7/1/2021 to 10/31/2021 of the larger OSCAR study. A single physician then adjudicated these cases as IHCA or not IHCA, and cases adjudicated as not IHCA were flagged as false positives.

We evaluated binary classification performance of encounter-level IHCA using precision, recall, and F1 score. Precision is equivalent to positive predictive value (PPV) and measures the accuracy of predictions made by the LLM. Recall is equivalent to sensitivity and measures the LLM’ ability to successfully identify all cases. F1 score is the harmonic mean between precision and recall. We evaluated multiclass classification performance using a confusion matrix, with correct predictions appearing along the matrix diagonal ^17^.

## Data Availability

Prompts are available upon reasonable request.

## Code Availability

Code available upon reasonable request.

## Author Contribution

J.V., D.W., K.R., S.C.C, and G.G.H. conceptualized and designed the study. J.V. and D.W. led the study. J.V., H.C., A.U.E., and K.O. collected and adjudicated the data. J.V. and D.W. created the models and performed data analysis. K.R., H.C., S.C.C, and G.G.H provided critical review of experiments. J.V., D.W., S.C.C, G.G.H, and K.R. contribute to the manuscript.

## Competing Interests

The authors declare no competing interests.

## Funding

No funding was used to support this research.

## Notes

### Competing Interest Statement

The authors have declared no competing interest.

### Funding Statement

This study did not receive funding.

### Author Declarations

The study was approved by the Cedars-Sinai Medical Center Institutional Review Board.

